# Deep learning-based Helicobacter pylori detection for histopathology: A diagnostic study

**DOI:** 10.1101/2020.08.23.20179010

**Authors:** Sharon Zhou, Henrik Marklund, Ondrej Blaha, Manisha Desai, Brock Martin, David Bingham, Gerald J. Berry, Ellen Gomulia, Andrew Y. Ng, Jeanne Shen

## Abstract

**Aims:** Deep learning (DL), a sub-area of artificial intelligence, has demonstrated great promise at automating diagnostic tasks in pathology, yet its translation into clinical settings has been slow. Few studies have examined its impact on pathologist performance, when embedded into clinical workflows. The identification of *H. pylori* on H&E stain is a tedious, imprecise task which might benefit from DL assistance. Here, we developed a DL assistant for diagnosing *H. pylori* in gastric biopsies and tested its impact on pathologist diagnostic accuracy and turnaround time.

**Methods and results:** H&E-stained whole-slide images (WSI) of 303 gastric biopsies with ground truth confirmation by immunohistochemistry formed the study dataset; 47 and 126 WSI were respectively used to train and optimize our DL assistant to detect *H. pylori*, and 130 were used in a clinical experiment in which 3 experienced GI pathologists reviewed the same test set with and without assistance. On the test set, the assistant achieved high performance, with a WSI-level area-under-the-receiver-operating-characteristic curve (AUROC) of 0.965 (95% CI 0.934–0.987). On *H. pylori*-positive cases, assisted diagnoses were faster (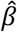, the fixed effect size for assistance = –0.557, p = 0.003) and much more accurate (OR = 13.37, p< 0.001) than unassisted diagnoses. However, assistance increased diagnostic uncertainty on *H. pylori-* negative cases, resulting in an overall decrease in assisted accuracy (OR = 0.435, p = 0.016) and negligible impact on overall turnaround time (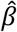 for assistance = 0.010, p = 0.860).

**Conclusions:** DL can assist pathologists with *H. pylori* diagnosis, but its integration into clinical workflows requires optimization to mitigate diagnostic uncertainty as a potential consequence of assistance.

## Introduction

*Helicobacter pylori* is the most prevalent chronic bacterial infection worldwide, affecting an estimated 4.4 billion individuals.^1^ A well-established association exists between chronic *H*. *pylori* infection and peptic ulcer disease, gastric cancer, and other gastric pathologies, as well as evidence linking infection to iron deficiency anemia, colorectal cancer, and other extra-gastric pathologies.^2–4^ Identifying and eradicating *H. pylori* in infected individuals reduces the risk of progression to long-term complications.^5,6^ Every gastric biopsy received in the pathology lab is evaluated for *H. pylori*, with the diagnosis resting on identification of one or more organisms upon high-magnification examination. Although the bacteria are readily identifiable on H&E stain in cases where high numbers of organisms are present, in other cases, making a diagnosis can be time consuming, tedious, and subject to interobserver variability, with missed diagnoses not uncommon.^7,8^ Although ancillary stains are available, these add significant cost and turnaround time to diagnosis,^8,9^ are not recommended for reflex application,^10,11^ and may be unavailable in low-resource settings. *H. pylori* diagnosis on H&E stain provides an ideal opportunity for deep learning (DL) assistance, which has already shown promise at automating other pathology tasks.^12-16^

Although DL (and other AI) models have demonstrated good performance on several histopathologic tasks, most studies have retrospectively compared model performance to that of a mixed group of diagnosticians of varying expertise levels. This may exaggerate the relative performance and clinical utility of the model, as the true end-users may be different (often with higher baseline diagnostic performance) from those against whom the model’s performance was compared. Furthermore, few studies have taken the next step of incorporating a model into a clinical workflow and evaluating its impact on users.^17,18^

In this study, we developed a DL ensemble of convolutional neural networks (CNNs) to assist pathologists with *H. pylori* diagnosis on H&E-stained whole-slide images (WSI), and sought to address the preceding gaps by testing the ensemble’s impact on the diagnostic performance of subspecialty GI pathologists.

## Materials and Methods

Institutional Review Board approval was obtained (IRB #48684), with waived informed consent for use of all patient material and data. The Standards for Reporting of Diagnostic Accuracy Studies (STARD)^19^ guidelines were used.

### Dataset and reference standard annotations

The study dataset consisted of H&E WSI of 311 gastric biopsies from 245 patients (160 *H*. *pylori*-positive and 151 *H. pylori*-negative biopsies), all with diagnostic confirmation by both H&E and *H. pylori* immunohistochemical (IHC) evaluation, obtained through stratified random sampling (maintaining an approximate 50:50 class balance of *H. pylori*-positive and –negative biopsies, for adequate representation of the morphologic range of each class) of all gastric biopsies submitted to our institution from January 1, 2015-December 31, 2018. All WSI were scanned at 40× magnification (0.25 micrometers per pixel) on an Aperio AT2 scanner (Leica Biosystems, Germany).

As WSI are too large to directly input into CNNs, they are subdivided into smaller image patches for input. Because not every patch in an *H. pylori*-positive WSI necessarily contains *H. pylori*, reference standard patch-level annotations were generated using *H. pylori* IHC. A sequential H&E de-stain/immunostain procedure was performed on the same tissue section to obtain annotations for the *H. pylori*-positive biopsies (see Figure 1a and Supplementary methods for details). This was done instead of using the original IHC slides, which contained sections cut at different depths within the block from the H&E slides, resulting in differences in the tissue and *H*. *pylori* content of the H&E and IHC slides (precluding accurate tissue co-registration and generation of reference standard annotations). For *H. pylori*-negative biopsies, an original diagnostic H&E slide was scanned from each biopsy.

**Figure 1.**
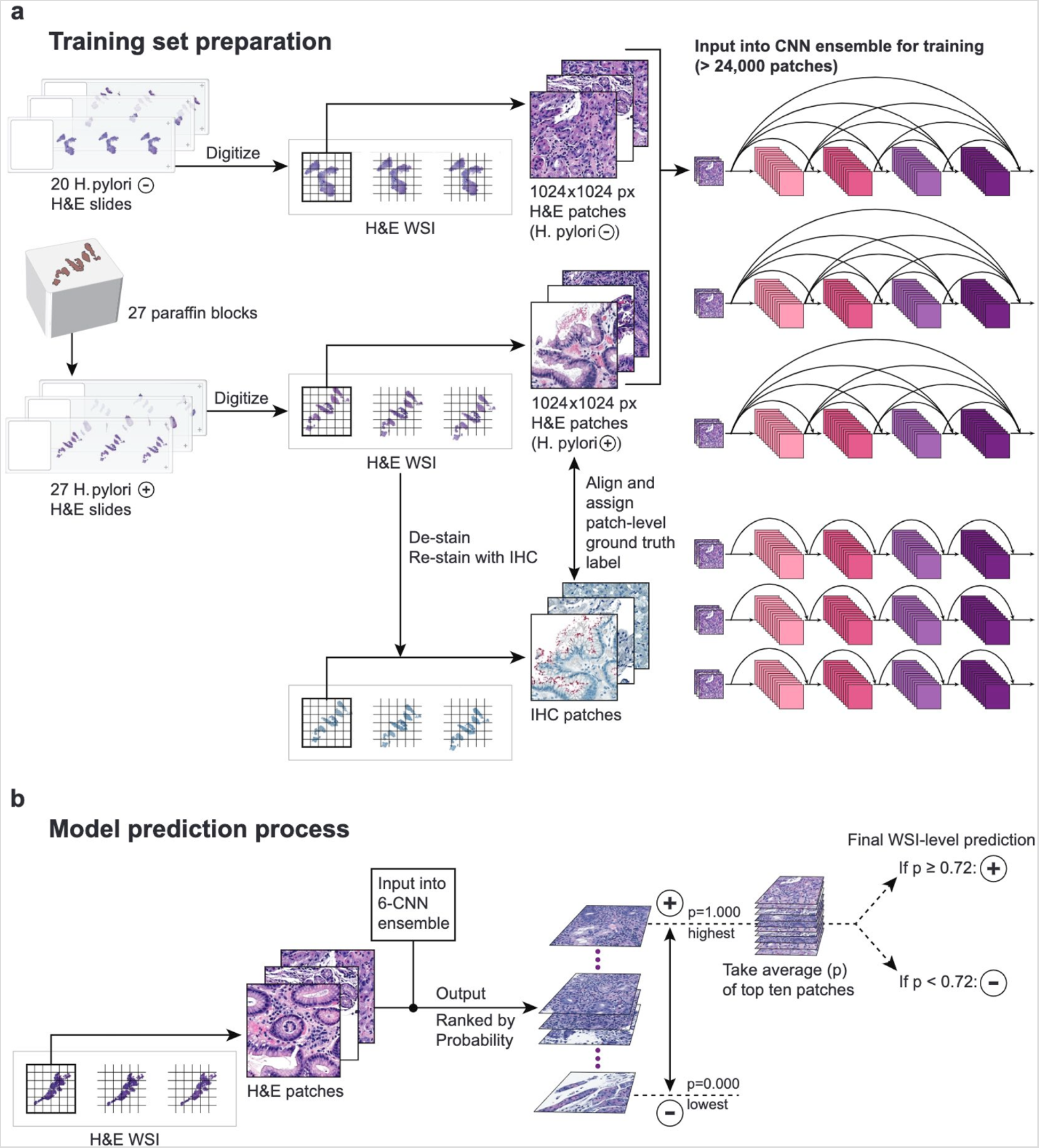
Training dataset preparation and model prediction process. **a,** The training dataset consisted of 24,786 non-overlapping image patches of size 1024 × 1024 pixels, extracted from one serial tissue section per H&E-stained slide (20 *H. pylori* negative and 27 *H. pylori* positive slides), input into a convolutional neural network (CNN) ensemble of 3 DenseNet-121 (top) and 3 ResNet-18 (bottom) architectures. To generate patch-level ground truth labels for the 27 *H. pylori*-positive slides, an H&E-stained slide was prepared from the paraffin block and digitized, then de-stained, re-stained with an *H. pylori* immunohistochemical (IHC) stain, and digitized to generate a paired IHC WSI for tissue co-registration with the H&E WSI. **b**, The ensemble’s WSI-level prediction of *H. pylori* status involved extracting all tissue-containing patches from a single level for input, computing the average of the patch-level probabilities for the top 10 highest probability patches, and binarizing this with a threshold of 0.72.

During the de-stain/re-stain process, eight slides experienced focal tissue detachment and/or incomplete immunostaining; these were excluded, resulting in a final dataset of 303 H&E WSI (152 *H. pylori*-positive and 151 *H. pylori*-negative), which was grouped by patient, shuffled, and randomly split (maintaining an approximate 50:50 positive:negative balance within each set) into a training set of 47 WSI (27 positive and 20 negative), validation set of 126 WSI (60 positive and 66 negative) for internal validation and optimization, and test set of 130 WSI (65 positive and 65 negative), which was completely held out from model training and internal validation/optimization, and used for the pathologist experiment. All WSI from the same patient were assigned to the same set. Each WSI was subdivided into non-overlapping image patches of size 1024 × 1024 pixels (256 × 256 micrometers) for model input, yielding, on average, 486 patches per WSI, and a total sample size of 147,258 patches. As multiple serial sections cut from the same block might be present on a WSI, only one section per WSI (the first, or left-most one on the slide) was used for training, yielding a training set of 24,786 patches. The 1024 × 1024 pixel patch size was chosen to be large enough to provide background tissue context for the models to learn from, but small enough for pathologists to easily view the entire patch at maximum resolution (40× magnification).

### Model development

Two CNN architectures, ResNet^20^ and DenseNet^21^, both of which have excelled on image classification benchmarks, were leveraged by averaging their predictions in a model ensemble, or set of CNNs whose outputs are combined to form a single prediction. Ensembling enables more robust prediction that is less sensitive to outlier predictions, as the final prediction comes, not from a single network, but from a collection of networks. Our ensemble consisted of three ResNet-18 and three DenseNet-121 architectures, each of which input an image patch and output a probability of *H. pylori* being present (for ensemble details, see Supplementary methods).

Patch-level probabilities were aggregated into WSI-level probabilities by averaging the top 10 patch-level probabilities from a single serial section (the first section) per WSI. As each WSI contained between 1–5 sections, only one section per WSI was used to avoid potential prediction bias related to the number of sections on a slide. The patch number (10) was determined empirically, based on the area under the receiver-operating-characteristic curve (AUROC) performance of the ensemble on the validation set. This 10-patch probability average was binarized using an optimal slide-level probability threshold (0.72) empirically determined from the ensemble’s performance on the validation set.

### Pathologist experiment

In a diagnostic study designed to simulate the workload in a high-volume pathology practice, we evaluated the ensemble’s impact on the accuracy and turnaround time of three subspecialty GI pathologists with 8, 9, and 27 years of respective practice experience, who reviewed the same test set (65 *H. pylori* positive and 65 *H. pylori* negative biopsies) during a single session, without time constraint. Half of each subgroup (positive and negative) was randomly assigned to review with ensemble assistance, while the remaining half was reviewed unassisted. The WSI sequence and assistance status were randomized for each pathologist (Figure 2a). The pathologists were blinded to all patient identifying and clinical information.

**Figure 2.**
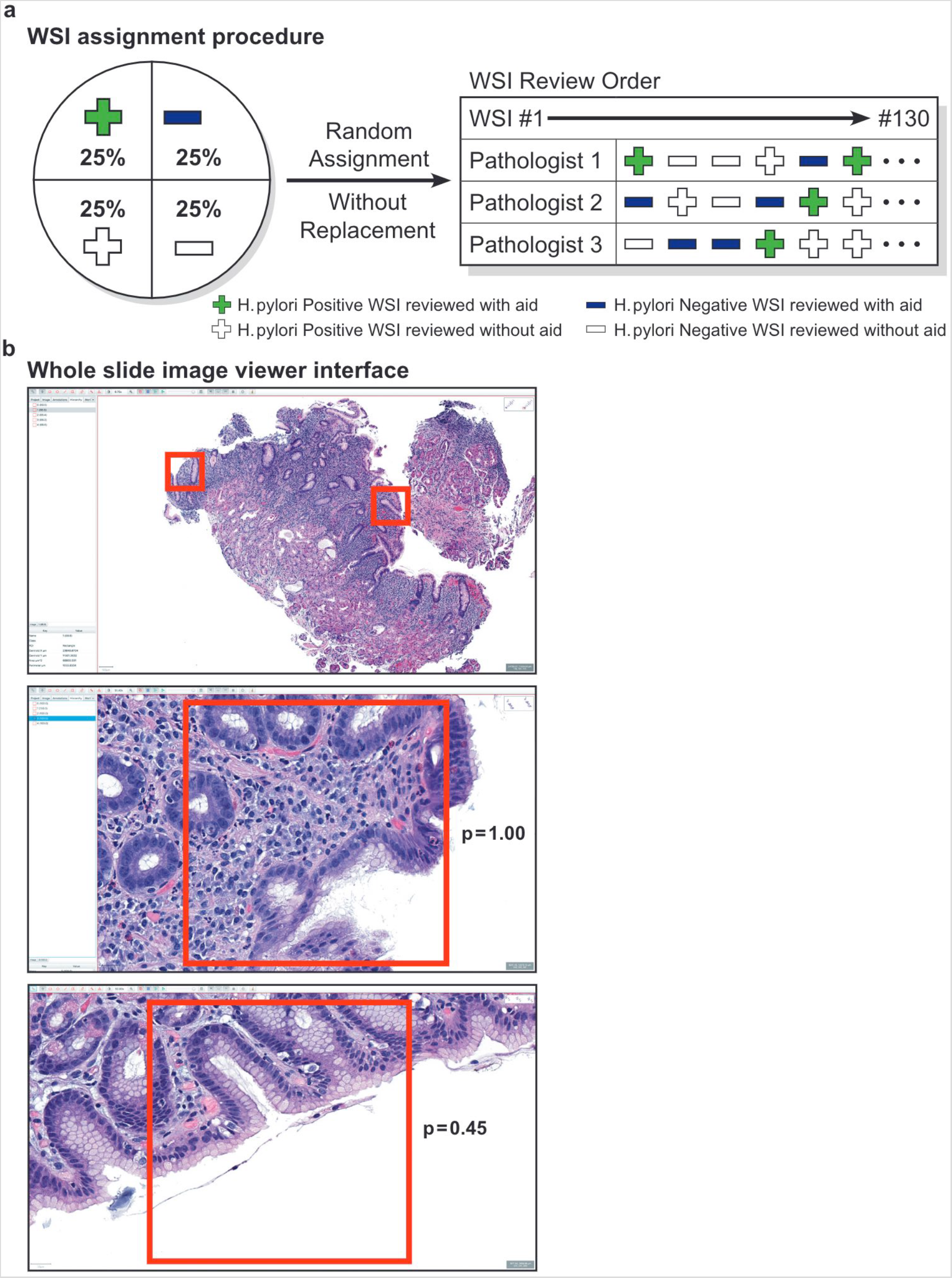
Design of the pathologist experiment and user interface. **a,** The 3 pathologists reviewed the same test set of 130 WSI (containing 65 positive and 65 negative WSI) during a single session, without time constraint. Half of the positive and negative WSI were randomly assigned to be reviewed with model assistance, with the remaining half assigned to be reviewed without assistance. Each pathologist received a unique randomized WSI review sequence and assignment of WSI to be reviewed with or without assistance. **b**, The WSI viewer interface consisted of a main WSI-viewing window (showing red bounding boxes around patches most likely to contain *H. pylori*) and a smaller window to the left displaying the list of all bounding boxes for that WSI, with their corresponding probabilities. By clicking on a bounding box in the smaller window, the user could automatically navigate to the corresponding region of the WSI in the main WSI-viewing window. The top panel shows a low magnification view, while the middle and bottom panels show higher magnification views with corresponding probabilities for two different bounding boxes from the same WSI. On the assisted cases, only the 3 to 5 bounding boxes that were most likely to contain *H. pylori* were displayed for each WSI. On unassisted cases, no bounding boxes or probabilities were displayed.

QuPath^22^, an open-source digital pathology package, was used for WSI review. Prior to the experiment, the ensemble was run on the 130 test WSI to generate patch-level *H. pylori* probabilities for every tissue-containing patch across all sections in each WSI, with the 3–5 highest-probability patches from each WSI and their corresponding probabilities selected for display. To avoid biasing the pathologists, binarized model predictions and the probability threshold for positivity were not shown. The ensemble made predictions on all sections present on the slide, reflective of a real-world practice scenario in which pathologists review all sections on a slide. (However, for the purposes of evaluating machine-learning performance metrics for the ensemble, only a single section per WSI was used.) For WSI containing only one section, bounding boxes for the 3 top-ranked patches were displayed. For WSI containing two or more sections, bounding boxes for the 5 top-ranked patches were displayed. This allowed the ensemble to make predictions across all tissue present in a WSI, while displaying a relatively limited number of patches, to avoid substantially slowing down the pathologists’ slide review.

The ensemble’s outputs were incorporated into the QuPath interface as follows: a window adjacent to the main WSI-viewing window displayed the list of all bounding boxes for a given WSI, with corresponding patch-level *H. pylori* probabilities. By clicking on a particular bounding box, pathologists could jump directly to the corresponding region in the main WSI-viewing window (see Figure 2b for an example). A numbered list of all 130 WSI was visible to the left of the main WSI-viewing window. WSI assigned for assisted review were marked with a text indicator (“Hierarchy”). On these cases, the pathologists were given discretion to review as few, or as many, of the bounding boxes and probabilities as they felt were necessary (including none). For WSI assigned to unassisted review, no bounding boxes or probabilities were displayed, and the pathologists simply navigated the WSI on their own.

For each WSI, the pathologist entered one of four diagnoses into a data entry software application which recorded a timestamp for computation of the diagnostic turnaround time: Positive (P) = *H. pylori* evident on H&E stain, Uncertain Positive (UP) = features suggestive of *H. pylori* infection, but would order IHC for confirmation, Uncertain Negative (UN) = most likely negative for *H. pylori*, but would order IHC for confirmation, and Negative (N) = would confidently diagnose as *H. pylori* negative on H&E stain. Use of the uncertain diagnostic choices (UP/UN) was intended to reflect clinical practice, where, rather than making an immediate choice between P or N, pathologists can be uncertain (resulting in a request for ancillary stains). Diagnoses were binarized for statistical analysis as follows: P and N diagnoses concordant with the reference standard annotation were considered correct, whereas UP and UN diagnoses, and P and N diagnoses discordant with the reference standard, were treated as incorrect. The rationale for treating uncertain diagnoses as incorrect was that uncertainty results in the performance of ancillary stains, which increases diagnostic turnaround time and cost.

The experiment was administered to each pathologist by the same administrator, who logged the time and duration of any interruptions (used to adjust the timestamp data for accurate determination of the per-slide turnaround time). All experiments were performed using the same workstation setup. Participants were given time at the beginning of each experiment to review a tutorial on use of QuPath and the data entry software application, and to practice the experiment workflow with 6 practice WSI that were not part of the 130 WSI test set.

### Statistical Analyses

We evaluated both the WSI-level and patch-level diagnostic performance of our ensemble using the AUROC, precision (positive predictive value), recall (sensitivity), specificity, accuracy, and F1-score, based on the respective probability binarization thresholds of 0.504 and 0.72 for patch and WSI-level predictions. Corresponding 95% confidence intervals for these metrics were calculated by bootstrapping, with a replicate size of 2,000.

Patch-level model performance was assessed on 1024 × 1024 pixel patches sampled from 10 *H. pylori*-positive and 10 *H. pylori*-negative WSI randomly selected from the 130 WSI test set. From each WSI, 100 patches were randomly sampled from a single section. If a WSI contained fewer than 100 patches in a section, all patches from that section were sampled. This resulted in a total of 871 patches from the 10 positive and 963 patches from the 10 negative WSI. Patch-level reference standard diagnoses for positive cases were obtained using the de-stain/re-stain method detailed previously, with confirmation by the reference pathologist. After excluding 87 patches with equivocal *H. pylori* status upon reference pathologist review, a total of 1,747 patches were used for patch-level performance evaluation.

The ensemble’s WSI-level performance was evaluated on all 130 WSI in the test set, where the average probability of *H. pylori* positivity across the 10 highest-probability patches from one section per WSI was binarized using the probability threshold of 0.72 WSI (Figure 1b). The same metrics used to evaluate patch-level performance were also calculated for WSI-level performance.

Pathologist performance was reported using the diagnostic accuracy (with 95% Wilson score confidence intervals^23^) and per-slide turnaround time (with 95% t-score confidence intervals). Our first objective was to investigate whether assistance was effective at increasing accuracy, based on the definitions of correct and incorrect diagnoses used to binarize the pathologist diagnoses. A generalized linear mixed model (GLMM) which included the assistance status (with or without assistance) and pathologist as fixed effects, and WSI as a random effect, was applied. The main effect of assistance was evaluated using a Wald z-test.

Our second objective was to investigate whether assistance affected the amount of time spent reaching the diagnosis. A log-normal mixed effect model, which included the assistance status and pathologist as fixed effects, and WSI as a random effect, was applied to estimate the effect of these variables on diagnostic turnaround time. The main effect of assistance was evaluated using a Wald t-test.

A significance level of α = 0.05 (two-tailed) was used for all statistical tests. The mixed effect models were developed using the Ime4^24^ and MASS^25^ packages in R.

## Results

### Model performance

On the validation set (126 WSI), our ensemble achieved a WSI-level AUROC = 0.952 (95% CI 0.913–0.983), with precision = 0.873 (95% CI 0.788–0.953), recall = 0.917 (95% CI 0.844–0.982), specificity = 0.880 (95% CI 0.794–0.955), accuracy = 0.897 (95% CI 0.841–0.944), and F1- score = 0.894 (95% CI 0.833–0.947). On the test set (130 WSI), the WSI-level AUROC = 0.965 (95% CI 0.934–0.987), with precision = 0.919 (95% CI 0.846–0.983), recall = 0.877 (95% CI 0.788-0.948), specificity = 0.924 (95% CI 0.857–0.984), accuracy = 0.900 (95% CI 0.846–0.946), and F1- score = 0.898 (95% CI 0.837–0.947). On the validation set, patch-level performance metrics were: AUROC = 0.877 (95% CI 0.863–0.881), accuracy = 0.872 (95% CI 0.862–0.881), F1 = 0.604 (95% CI 0.574–0.633), precision = 0.616 (95% CI 0.580–0.651), recall = 0.593 (95% CI 0.558-0.629), and specificity = 0.927 (95% CI 0.918–0.935), using a probability threshold of 0.504 for *H*. *pylori* positivity. On the test set, the patch-level AUROC = 0.911 (95% CI 0.888–0.933), with accuracy = 0.892 (95% CI 0.878–0.907), F1 = 0.573 (95% CI 0.516–0.627), precision = 0.488 (95% CI 0.428–0.549), recall = 0.692 (95% CI 0.623–0.760), and specificity = 0.916 (95% CI 0.902-0.929).

### Model impact on pathologist performance

During the pathologist study, four reads were excluded due to data entry errors made by the pathologists while recording diagnoses into the software application, resulting in a final set of 386 pathologist reads for analysis. The overall accuracy of the pathologists was 0.680 (95% CI 0.611–0.742) on unassisted cases and 0.573 (95% CI 0.502–0.641) on assisted cases. After controlling for pathologist and WSI effects, we found that assistance had a significant positive impact on *H. pylori*-positive cases, with assisted diagnoses being more accurate (OR = 13.37, 95% CI 3.622–49.320, p< 0.001) than unassisted diagnoses. However, on *H. pylori*-negative cases, assistance had a negative impact, with unassisted diagnoses being 2.30 times more likely to be correct than assisted diagnoses (OR for correct diagnosis = 0.435, 95% CI, 0.215-0.844 p = 0.016), resulting in a decrease in overall accuracy (Figure 3b).

**Figure 3.**
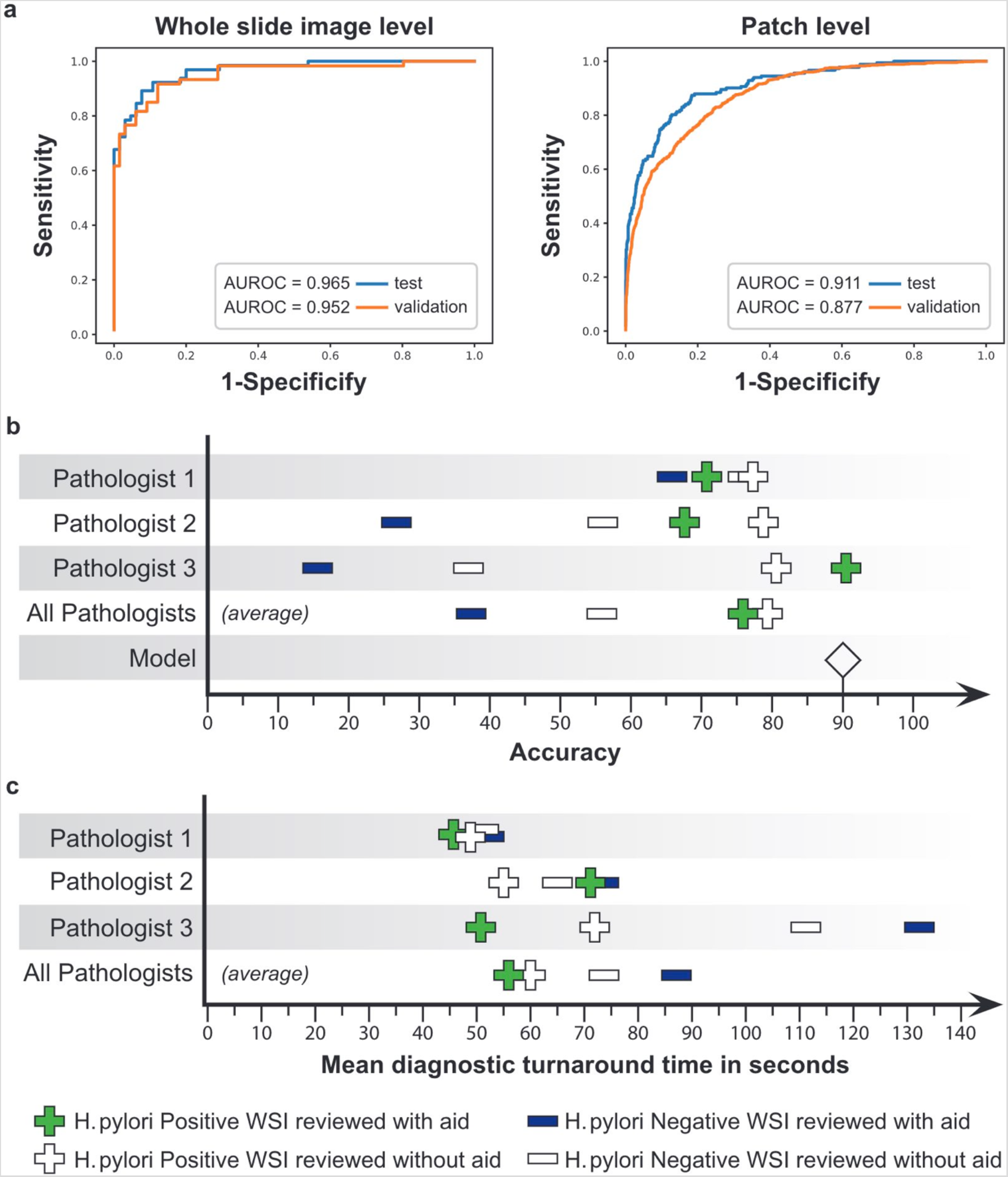
Performance of the model ensemble and impact of assistance on pathologist accuracy and diagnostic turnaround time. **a,** The WSI (left) and patch (right) level receiver-operating-characteristic (ROC) curves for the best-performing model ensemble on the validation (orange, n = 126 WSI) and test (blue, n = 130 WSI) datasets are shown, along with respective area under the curve (AUROC). **b**, The assisted and unassisted pathologist accuracies on *H. pylori* positive and negative WSI in the 130 WSI test set are shown. On *H. pylori*-positive WSI, accuracies for individual pathologists ranged from 0.774–0.818 without assistance, and 0.688–0.906 with assistance. On *H. pylori* negative WSI, individual pathologist accuracies ranged from 0.375–0.758 unassisted and 0.161–0.677 assisted. The accuracy of the DL ensemble alone on the test set was 0.900. **c**, The average per-WSI diagnostic turnaround times (TAT) with and without assistance are shown. On *H. pylori*-positive WSI, individual pathologist TAT ranged from 49.7–73.7 seconds unassisted and 46.1–71.6 seconds assisted. On *H. pylori* negative WSI, individual TAT ranged from 47.8–110.8 seconds unassisted and 50.9–133.0 seconds assisted.

The average per-slide turnaround time was 67.15 seconds (95% CI 59.11–75.20 s, range 6–408 s) in the unassisted state, and 70.69 seconds (95% CI 63.00–78.38 s, range 11–356 s) in the assisted state. The average per-slide turnaround time for each diagnostic category was: P = 43.16 seconds (95% CI 35.60–50.73 s, range 6–356 s), N = 51.87 seconds (95% CI 46.53-57.21s, range 16–168 s), UP = 129.68 seconds (95% CI 115.30–144.07 s, range 47–408 s), and UN = 84.13 seconds (95% CI 75.74–92.53 s, range 39–221 s) (Figure 4a). After controlling for pathologist and WSI effects, we found a significant reduction in diagnostic turnaround time for *H. pylori-* positive cases (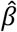, the fixed effect size for assistance = –0.557, 95% CI –0.338-0.119, p = 0.003), which was offset by a slower average turnaround time on negative cases, resulting in an overall negligible change in turnaround time with assistance (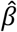=0.010, 95% CI –0.097–0.116, p = 0.860).

**Figure 4.**
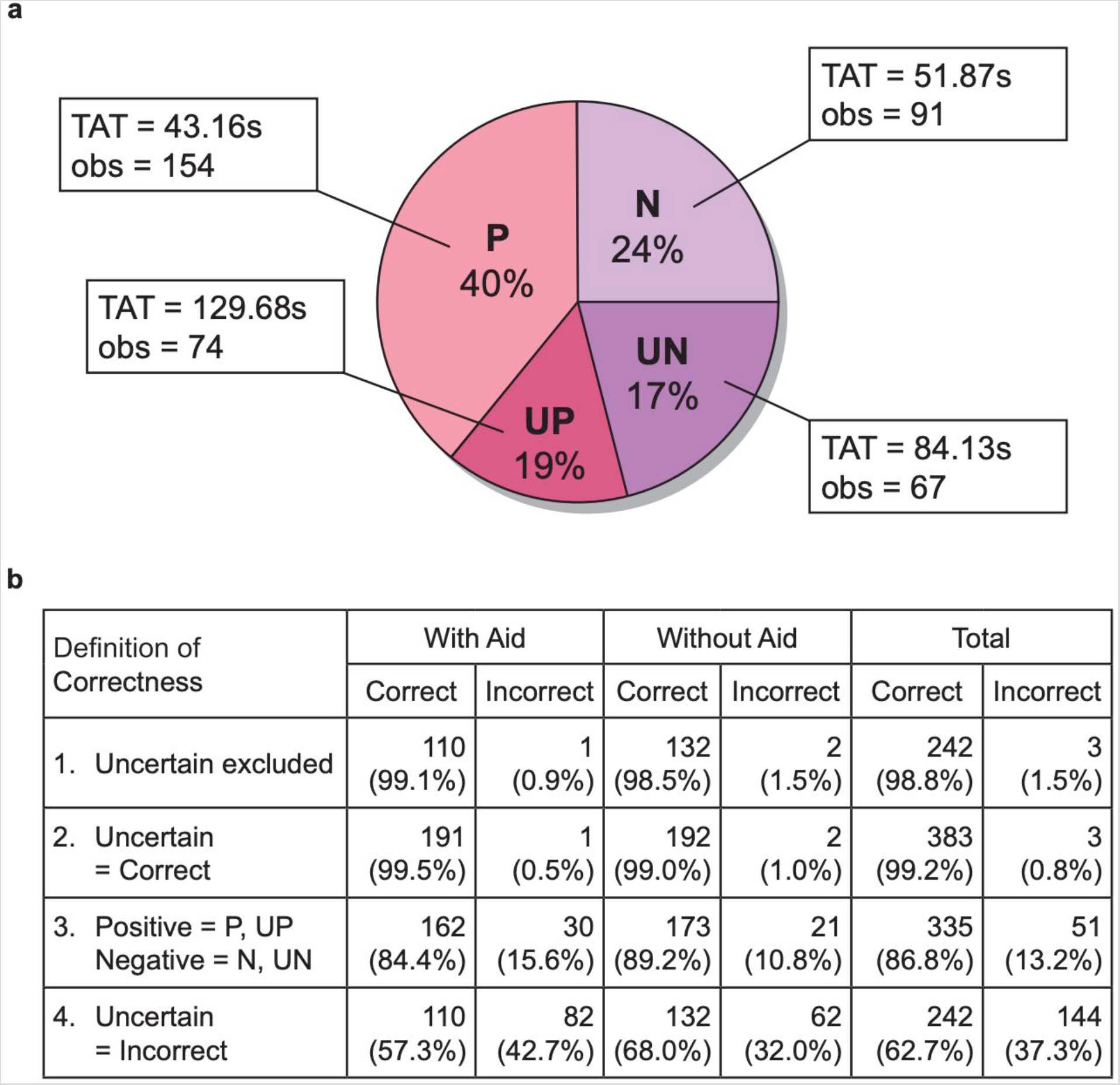
Results of the pathologist experiment. **a,** The number and percentage of total observations (diagnoses made by the pathologists) and turnaround time (TAT) in seconds are shown for each diagnostic category, where P = Positive, N = Negative, UP = Uncertain Positive, UN = Uncertain Negative. **b**, The numbers and percentages of correct and incorrect diagnoses made by the pathologists, given different possible definitions of correctness, are shown. For the analyses performed in the study, the last definition of correctness, where all Uncertain diagnoses were considered incorrect, was used.

## Discussion

In this study, we evaluated the impact of DL assistance on the diagnostic accuracy and turnaround time of subspecialty GI pathologists, for the routine task of diagnosing *H. pylori* on H&E-stained gastric biopsies. We observed a significant improvement in both metrics with assistance on *H. pylori*-positive cases, but a detrimental effect of assistance on both metrics for *H. pylori*-negative cases.

In particular, the negative impact of assistance when evaluating *H. pylori*-negative cases resulted in a decrease in overall pathologist accuracy, which might be explained by increased diagnostic uncertainty with assistance, with 81 versus 60 uncertain diagnoses made in the assisted and unassisted states, respectively. Because the pathologists were not informed of the probability threshold for positivity (to avoid biasing their diagnoses), subjectivity in their interpretation of the probabilities could have contributed to increased uncertainty. Based on the method used to binarize diagnoses into correct and incorrect categories (uncertain diagnoses counted as incorrect, Figure 4b), 20 more incorrect diagnoses were made with assistance. However, when uncertain diagnoses were excluded from analysis, more correct diagnoses were made with, versus without, assistance, suggesting that the detrimental effect of assistance was primarily attributable to increased diagnostic uncertainty, rather than to increased diagnostic error. Increased uncertainty may be an unintended consequence of AI assistance, which should be considered when designing and incorporating AI models into clinical practice. For example, a follow-up study of the current model might evaluate the impact of assistance when binarized outputs are displayed, rather than probabilities.

During the experiment, the pathologists were provided with the 3–5 top-ranked patches on each assisted case, regardless of what the corresponding patch-level probabilities were. Although they were allowed to review these patches at their discretion, they might have felt obligated to view at least some, or even all, patches, simply because these were being presented. This might have introduced diagnostic uncertainty where there initially was not, contributing to additional turnaround time. Given the significant benefit of assistance on *H. pylori*-positive slides, future user interfaces might be designed so that assistance is provided only when patches exceed a probability threshold for positivity, and model outputs are hidden when a slide is predicted to be negative.

While many studies of medical AI models have emphasized diagnostic accuracy, sensitivity, or specificity as primary metrics, less attention has been devoted to examining the impact on other practical considerations, such as turnaround time. In our study, the average per-slide turnaround time was approximately 67 seconds unassisted and 71 seconds assisted, with no statistically significant difference in turnaround time with assistance, after controlling for pathologist and WSI effects. Given these relatively quick turnaround times, the impact of assistance on this metric might be considered inconsequential, from a clinical standpoint. However, the potential for human-computer interaction to increase turnaround time in prospective diagnostic settings should be highlighted, as it may cause unintended harm to patients when occurring in situations where diagnostic turnaround time is of utmost importance. For the current application, the longer turnaround times for uncertain diagnoses (approximately 130 and 84 seconds for UP and UN diagnoses, respectively) suggest that a decrease in overall turnaround time could be reached by reducing uncertainty in human-AI interaction during clinical workflow integration.

Potential solutions for mitigating uncertainty might be to present the binarization threshold used for *H. pylori* positivity, to display explicitly binarized model outputs, or, as previously discussed, to set a probability threshold for presenting pathologists with model outputs. Another solution might be to deploy the model, not as a primary diagnostic assistant, but as an automated pre-screening tool, given its fast processing time, ability to correctly diagnose *H. pylori* on slides where the bacteria are present, and lack of human diagnostic uncertainty. The model’s accuracy on the test set was 90%. In contrast, the pathologists’ unassisted accuracy was 89.2%, when P/UP diagnoses were counted as positive and N/UN diagnoses counted as negative (the closest post-hoc approximation of a “no-uncertainty” scenario). The reported pathologist sensitivity and specificity of *H. pylori* diagnosis on H&E stain ranges from 69–93% and 87–90%, respectively.^8^ Our ensemble achieved a comparatively good sensitivity of 87.7% and a specificity of 92.0% (higher than the upper range of reported pathologist specificities). If a different operating point on the ensemble’s ROC curve were selected to further maximize the sensitivity while retaining acceptable specificity, cases flagged as negative by the ensemble might be shifted to the end of the pathologist queue, while those flagged as positive might be prioritized for review. Negative cases would no longer need to be painstakingly reviewed for *H*. *pylori* or submitted for ancillary testing, while positive cases could be reviewed with the top-ranked patches shown first, potentially reducing diagnostic turnaround time and ancillary staining costs. Yet another deployment option might be to incorporate the ensemble into an automated “double-check” tool, which could run in the background and alert the pathologist of any disagreement between their diagnosis and the ensemble’s prediction.

In this study, we tested the impact of the ensemble on subspecialty GI pathologists, as this is the group that most commonly reviews gastric biopsies (and the only one that does so at our institution). Given their high baseline unassisted accuracy (when uncertain diagnoses were excluded, a total of only 2 incorrect diagnoses were made), it is somewhat expected that assistance did not result in a statistically significant improvement in accuracy. While we did not test the impact on non-GI pathologists, trainees, or other subgroups with less experience at the task, it is possible that our ensemble could significantly improve accuracy and turnaround time for these other subgroups. In practices where there is a shortage of GI-trained pathologists, models such as the one in this study might provide value as a pre-screening or primary diagnostic tool. To our knowledge, this is the first study to develop and test the impact of an AI model for the direct detection of *H. pylori* organisms on H&E-stained whole-slide images.

Our study was subject to limitations. While the ensemble’s accuracy was dependent on the selected binarization threshold for *H. pylori* positivity, the pathologists’ accuracy was dependent on binarization of four possible diagnoses into correct and incorrect categories. Use of the UP and UN categories precluded direct comparison of ensemble performance with that of the pathologists. In theory, a direct comparison might be made if the ensemble’s outputs were thresholded into the same diagnostic choices available to the pathologists. In reality, the degree of interobserver variability in the uncertainty threshold among different pathologists would make it nearly impossible to establish a reliable threshold for the uncertain categories (whereas *H*. *pylori-*positive and –negative diagnoses have a ground truth, there is none for uncertain diagnoses).

Our ensemble was developed to help pathologists identify the presence of *H. pylori*, an essential task which is performed, without exception, on every gastric biopsy. We acknowledge that other concurrent pathologies may be present in the same biopsy, which are not currently addressed by the ensemble, and which could be incorporated into future diagnostic suites meant to assist with general gastric biopsy review.

Finally, our study was limited to data from a single pathology department serving a regional healthcare system, with an imposed 50:50 class balance of positive and negative cases (chosen to obtain broad representation of the morphologic range of cases, but which also reflects the worldwide prevalence of *H. pylori* infection^1^). Future studies incorporating datasets from multiple institutions and regions, as well as more pathologists, are recommended to validate our findings.

### Conclusions

DL can diagnose *H. pylori* in H&E-stained gastric biopsies with high performance, and holds potential for automating this common diagnostic task. However, contrary to prevailing expectations regarding AI assistance, even a DL model with good performance metrics may fail to improve human diagnostic performance, if it is not integrated into clinical workflows in an optimal way. Although AI holds promise for improving healthcare quality and efficiency, its ultimate clinical impact may be determined, not by model performance metrics, but by the manner in which clinicians interact with these models. Our results suggest that increased diagnostic uncertainty is an important unintended consequence of human-AI interaction, which may decrease diagnostic accuracy and lead to longer case turnaround times. We hope that our findings present a realistic picture of AI’s impact on pathologists, while encouraging greater attention toward addressing the various aspects of human-computer interaction that will determine the ultimate real-world impact of these models.

## Data Availability

The whole-slide images used in the study are not currently publicly available, in accordance with institutional requirements governing human subject privacy protections.

## Author Contributions

### Concept and design

Zhou, Marklund, Ng, Shen

### Acquisition, analysis, or interpretation of data

Zhou, Marklund, Blaha, Desai, Bingham, Martin, Berry, Gomulia, Shen

### Drafting of the manuscript

Zhou, Marklund, Blaha, Shen

### Manuscript revision and approval

Zhou, Marklund, Blaha, Desai, Bingham, Martin, Berry, Gomulia, Ng, Shen

### Statistical analysis

Zhou, Marklund, Blaha, Desai, Shen

### Administrative, technical, or material support

Zhou, Marklund, Gomulia, Ng, Shen

### Supervision

Ng, Shen

## Acknowledgments

We thank Norman L. Cyr (Graphic Arts & Imaging Services, Department of Pathology, Stanford University) for his assistance in creating the figures for this article. This study was supported by the Department of Pathology, Stanford University, with additional infrastructure provided by the Stanford Machine Learning Group and the Stanford Center for Artificial Intelligence in Medicine & Imaging.

## Notes

**Conflicts of interest**: The authors declare no competing interests.

### Competing Interest Statement

The authors have declared no competing interest.

### Funding Statement

No external funded was received for the work reported in this study.

### Author Declarations

The Stanford University IRB provided waived informed consent for this study.

## References

1. Hooi JKY, Lai WY, Ng WK, et al. Global Prevalence of Helicobacter pylori Infection: Systematic Review and Meta-Analysis. Gastroenterology. 2017;153(2):420–429.

2. Marshall BJ, Warren JR. Unidentified curved bacilli in the stomach of patients with gastritis and peptic ulceration. Lancet. 1984;1:1311–1315.

3. International Agency for Research on Cancer. Schistosomes, liver flukes and Helicobacter pylori, IARC Working Group on the Evaluation of Carcinogenic Risks to Humans, vol. 61. Lyon, France: IARC; 1994.

4. Crowe SE. Indications and diagnostic tests for Helicobacter pylori infection. In Grover, S. (Ed.), UpToDate. https://www.uptodate.com/contents/indications-and-diagnostic-tests-for-helicobacter-pylori-infection. Retrieved October 2, 2019

5. Malfertheiner P, Megraud F, O’Morain CA, et al. Management of Helicobacter pylori infection-the Maastricht V/Florence Consensus Report. Gut. 2017;66(1):6–30. doi: 10.1136/gutjnl-2016-312288.

6. Chey WD, Wong BC, Practice Parameters Committee of the American College of Gastroenterology. American College of Gastroenterology guideline on the management of Helicobacter pylori infection. Am J Gastroenterol. 2007;102(8):1808.

7. Faigel DO, Childs M, Furth EE, Alavi A, Metz DC. New noninvasive tests for Helicobacter pylori gastritis. Comparison with tissue-based gold standard. Digest Dis Sci. 1996;41(4):740–748.

8. Lee JY, Kim N. Diagnosis of Helicobacter pylori by invasive test: Histology. Ann Transl Med. 2015;3(1):10.

9. 2019 Medicare Physician Fee Schedule. https://documents.cap.org/documents/2019-final-medicare-impact-table_181106_10474.pdf. Retrieved Jan 1, 2019.

10. Batts KP, Ketover S, Kakar S, et al. Appropriate use of special stains for identifying Helicobacter pylori: Recommendations from the Rodger C. Haggitt Gastrointestinal Pathology Society. Am J Surg Pathol. 2013;37(11):e12–e22. doi:10.1097/PAS.0000000000000097

11. Pittman ME, Khararjian A, Wood LD, Montgomery EA, Voltaggio L. Prospective identification of Helicobacter pylori in routine gastric biopsies without reflex ancillary stains is cost-efficient for our health care system. Hum Pathol. 2016;58:90–96. doi:10.1016/j.humpath.2016.07.031

12. Salto-Tellez M, Maxwell P and Hamilton P. Artificial intelligence—the third revolution in pathology. Histopathology. 2019;74: 372–376.

13. Bejnordi BE, Veta M, van Diest J, et al. Diagnostic assessment of deep learning algorithms for detection of lymph node metastases in women with breast cancer. JAMA. 2017;318(22):2199–2210.

14. Strom P, Kartasalo K, Olsson H, et al. Artificial intelligence for diagnosis and grading of prostate cancer in biopsies: a population-based, diagnostic study. Lancet Oncol. 2020;21(2):222–232.

15. Veta M, Heng YJ, Stathonikos N, et al. Predicting breast tumor proliferation from whole-slide images: The TUPAC16 challenge. Med Image Anal. 2019;54:111–121.

16. Stalhammar G, Robertson S, Wedlund L, et al. Digital image analysis of Ki67 in hot spots is superior to both manual Ki67 and mitotic counts in breast cancer. Histopathology. 2018;72(6):974–989.

17. Steiner DF, MacDonald R, Liu Y, et al. Impact of deep learning assistance on the histopathologic review of lymph nodes for metastatic breast cancer. Am J Surg Pathol. 2018;42(12):1636–1646.

18. Kiani A, Uyumazturk B, Rajpurkar P, et al. Impact of a deep learning assistant on the histopathologic classification of liver cancer. NPJ Digital Med. 2020;3:23; doi:10.1038/s41746-020-0232-8

19. Bossuyt PM, Reitsma JB, Bruns DE, et al; STARD 2015: an updated list of essential items for reporting diagnostic accuracy studies. BMJ. 2015;351:h5527. doi:10.1136/bmj.h5527.

20. He K, Zhang X, Ren S, Sun J. Deep residual learning for image recognition. 2016 IEEE Conference on Computer Vision and Pattern Recognition (CVPR). 2016:770–778.

21. Huang G, Liu Z, van der Maaten L, Weinberger KQ. Densely connected convolutional networks. 2016: Preprint at https://arxiv.org/abs/1608.06993

22. Bankhead P, Loughrey MB, Fernandez JA, et al. QuPath: Open source software for digital pathology image analysis. Sci Rep. 2017;7(1):16878.

23. Wilson EB. Probable inference, the law of succession, and statistical inference. J AM Stat Assoc. 1927;22:209–212.

24. Bates D, Mächler M, Bolker B, Walker S. Fitting linear mixed-effects models using lme4. J Stat Softw. 2015;67:1–48.

25. Venables WN, Ripley BD. Modern Applied Statistics with S, Fourth edition. New York, NY:Springer;2002.

26. Martin DR, Hanson JA, Gullapalli RR, Schultz FA, Sethi A, Clark DP. A deep learning convolutional neural network can recognize common patterns of injury in gastric pathology. Arch Pathol Lab Med. 2020;144(3):370–378.

